# Specimen sharing for epidemic preparedness: building a local-to-global virtual biorepository framework

**DOI:** 10.1101/2023.01.17.23284659

**Authors:** Judith Giri, Laura Pezzi, Rodrigo Cachay, Rosa Margarita Gèlvez Ramirez, Adirana Tami, Sarah Bethencourt, Anylea Lozano, Julia Poje, Thomas Jaenisch, May Chu

## Abstract

We present a framework for a federated, virtual biorepository system (VBS) with locally collected and maintained specimens, based on a ‘global goods’ model and principles of equitable access and benefit sharing. The VBS is intended to facilitate timely access to biological specimens and associated data for outbreak-prone infectious diseases to accelerate the development and evaluation of diagnostics, assess vaccine efficacy, and to support surveillance and research needs. The VBS is aimed to be aligned with the WHO BioHub and other specimen sharing efforts as a force multiplier to meet the needs of strengthening global tools for countering epidemics. The purpose of our research is to lay the basis of the collaboration, management and principles of equitable sharing focused on low- and middle-income country partners. Here we report on surveys and interviews undertaken with biorepository-interested parties to better understand needs and barriers for specimen access and share examples from the ZIKAlliance partnership on the governance and operations of locally organized biorepositories.

## Introduction

The continuing and evolving emerging epidemics - Ebola, Zika and COVID-19 to name only a few - highlight the need for timely, transparent and efficient access to quality, annotated specimens to drive research, development of new diagnostics and treatments, disease surveillance, and to measure vaccine efficacy on a global scale. Accessing quality specimens has been largely driven by pathogen-specific needs and related research interests, with little consideration to the broader sharing of those specimens or collections. Trusted infrastructure and specimen sharing mechanisms need to be in place to support the needs of emergent outbreak responses. Without a supportive infrastructure, equitable and ethical sharing of specimens can become complicated, with layers of barriers that lead to prolonged and stressful processes, blunting the best intentions to quickly respond to public health emergencies [1-6].

Biorepositories (biobanks) retain and collect specimens (samples) as a fundamental resource for advancing research and serve as essential infrastructure for responsible and ethical specimen management. Biorepositories are also rapidly evolving and gaining more prominence in the data-driven health research arena [7-10]. Great strides have been made from the hand labeled small collection in an investigator’s freezer to the professionally managed institutional facilities, with quality management systems based on best practices [11] and accreditation standards-ISO 20387;20189(E) Biotechnology-Biobanking standards (https://www.iso.org/obp/ui/#iso:std:iso:20387:ed-1:v1:en) supported by robust inventory systems and associated clinical data. Most large-scale biorepositories have been established to meet specimens needs for non-communicable diseases like cancer or genetic disorders and, more recently, to benefit personalized medicine [9,11,12]. Large population-based public health and clinical biorepositories have also been established such as the United Kingdom Biobank (https://www.ukbiobank.ac.uk/), the National Health and Nutrition Examination Survey (https://www.cdc.gov/nchs/nhanes/biospecimens/biospecimens.htm), the National Cancer Institute (https://www.cancer.gov/research/infrastructure) as well as biobank hubs such as the German Biobank Node [13]. The COVID-19 pandemic has led to a proliferation of collections both in existing and COVID-19-centric biorepositories (PATH, https://www.path.org/programs/diagnostics/washington-covid-19-biorepository/ [14-16]. Yet, despite the existence of these developments there is still room for additional collections as these do not represent the diverse range of geography, ecozones, and demographics needed to support emerging infectious disease research response efforts. The need for broader specimen collections for (emerging) infectious diseases, representative of diverse populations, especially from LMIC, remains an unmet need - and even given recent investments during COVID-19, a transparent sharing system has not been fully realized [5,6,14,17].

A major challenge remains the coordination of biorepositories and collections in different geographic areas and overcoming obstacles for access. A few successful models such as the Foundation for Innovative Diagnostics (FIND) DxConnect (https://www.finddx.org/wp-content/uploads/2022/12/20211202_fac_specimen_bank_VF_EN.pdf) and the European Viral Archive-Global provides a trusted broker role in accessing virus strains, molecular targets, virus components and cell lines [2,18] and in this paper, the ZIKAlliance network of local biorepositories all showing that there are communities of practice willing to share and have needs to access timely, quality specimens.

A frequently invoked barrier is the benefit sharing requirement of the Convention for Biological Diversity’s Nagoya protocol (Protocol, The Nagoya Protocol on Access and Benefit-sharing (cbd.int)). The Nagoya protocol mandates signatories to the Protocol to comply with respective national requirements for benefits to be equitable and fairly shared if materials from that country contributed to the development of products such as diagnostic tests, vaccines, or therapeutics. National signatories have adopted differing interpretations of the Protocol which has become potential barriers for access to specimens [19]. The full impact and benefits enshrined in the Protocol remains a work in progress and is being actively addressed by the WHO Biohub effort.

A serious challenge in sourcing specimens across countries and institutions is the inherent heterogeneity of methods of collection, characterization and handling of specimens and data, and the difficulty this poses for users regarding the quality and integrity of the specimens. This is not surprising because maintenance of biorepositories is typically an unfunded mandate for a clinic or laboratory. Without active efforts to standardize, it is not surprising that infectious disease biorepositories are fragmented and pose challenges to the stakeholder community trying to create access to specimens. Fortunately, we have been aware of several functional efforts which can serve as models to guide our approaches for a harmonized VBS framework.

The abundance of specimens during the COVID-19 pandemic, unlike previous situations in outbreaks where competition for a limited number of specimens was the rule, provided a good opportunity to examine existing gaps for specimen collection, their use, and sharing mechanisms [1,5,16]. We identified that there are many untapped and interested sites that could be contributors and users of specimens, but lack a holistic and more cohesive approach to identify and tackle the barriers to receive the benefits of sharing. An approach, centered on participation of low-and-middle income countries (LMIC), who often are at the center of new public health threats or emerging zoonotic hot zones, could prove pivotal in enabling LMIC to lead collecting and managing specimens for broader sharing [3,20,21].

Therefore, we sought to seek input from a broad group with interest in accessing/providing specimens, especially for serological diagnostics, to examine how to best provide practical solutions that take advantage of existing collections rather than investing in building large new biorepository facilities. We initiated a series of workshops, and while heavily interrupted by the emergence and spread of COVID-19, the pandemic also provided an opportunity to engage many more who are interested in specimen sharing.

The efforts leading up to this manuscript were initiated at a workshop at ASTMH in 2019, co-convened by Center for Global Health (CGH) at the Colorado School of Public Health and FIND as an outcome of the 2016 Zika virus public health emergency. We have subsequently organized virtual workshops hosted by the Global Health Network (TGHN; https://globalbiorepository.tghn.org/) and initiated a survey and interviews with these goals in mind: to identify the current barriers to specimens sharing and unmet needs of various stakeholders; and initiate discussion about proposed distributed, locally-managed virtual biorepository system (VBS) as a solution to meeting key challenges.

We based our questions in the survey and the interviews on a vision for a VBS characterized by principles of equitable, transparent access to specimens for the ‘global good’, and at the same time responding to the needs of diverse stakeholders. We focused on diseases caused by pathogens with outbreak potential and placed emphasis on specimens for development of diagnostics and research. In our survey and interviews we asked participants what their view might be for a sustainable infrastructure focused at the LMIC level that provides benefits to diverse specimen consumers and providers.

We intend to use a grassroots approach to developing a VBS that will support local governance and stewardship of specimens. For that reason, we sought input from local decision-making and governance structures in LMICs regarding the sharing of biological material, incorporating longstanding collaborative relationships of the authors (e.g., within the Reconciliation of Cohort Data for Infectious Diseases (ReCoDID), www.recodid.eu; and ZIKAlliance consortia, https://zikalliance.tghn.org/ and AEDES Network, https://www.redaedes.org/). Since 2021, we have joined forces with the Center for Research in Emerging Infectious Disease (CREID) Coordinating Center (https://creid-network.org/coordinating-center) to guide the activities of the VBS.

## Methods

### Participation and privacy policy

Participants included in our workshops and survey demonstrate a broad representation of individuals who had expressed interest in biorepository activities (either through contact with our website hosted by the Global Health Network or from individuals who volunteered to join the discussions on a personal interest basis. Participants were informed that they retained the right to withdraw without penalty and the collected data would be de-identified and reported in aggregate form following the research-ethics guidelines of the American Psychological Society research-ethics guidelines (https://www.apa.org/ethics/code/ethics-code-2017.pdf). The registered IRB protocol waiver is #23-0079 from the University of Colorado-Anschutz.

### Consultative workshops and recruitment of participants

Workshops were conducted to identify what the challenges and needs might be to build a durable grassroots system to access specimens. Three workshops, in person (2019) and online conferences (2020 and 2021) were convened to identify, evolve and fine-tune the tasks for the VBS. Invitations were extended to those individuals and/or representatives of agencies and entities who had joined previous biorepository activities, to those identified through publications as well as those who had expressed interest through TGHN website. The scope began by looking at the challenges encountered during the ZIKV epidemic in South America and the Caribbean. Then the scope was extended to include discussion of the COVID-19 pandemic and outbreak-prone diseases in the second and third workshops to explore potential benefits on for both the sample contributor as well as the user or ‘client’ side.

### Survey on benefits in participating in a VBS

A self-administered 10-question qualitative survey (Supporting Information, SI; Table 1) was created on the SurveyMonkey platform for the participants. The survey was accessed by email or website (https://globalbiorepository.tghn.org/) between December 2020 through January 2022. Data analysis was finalized in December 2022.

### Interviews to assess needs and barriers to accessing specimens

Video-conference interviews were conducted jointly by 2 members of the study team (JG and MC) and recorded with the consent of the interviewees. We targeted potential users of well-characterized specimens, including commercial as well as academic entities. Commercial users consisted of diagnostics industry companies that represented both small in vitro diagnostics (IVD) products <10 and mid-size to larger (with IVD products > 10) operations [22]. We also interviewed not-for-profit commercial, research and academic/government entities with regard to their need and use of biological samples. Our questions were based on their respective strategies in developing diagnostic tests and reference controls and how they would acquire materials for developing an IVD. Interviewees were encouraged to provide additional feedback about specific hurdles they encountered sourcing specimens. The interviews lasted on average from 40 to 60 minutes and responses were compared and tabulated for analysis. The complete seven question survey is in SI, Table 2.

### Local biorepository governance structures

We also conducted interviews with research teams from the ZIKAlliance consortium [21] including: (a) the Industrial University of Santander, Bucamaranga, Colombia; (b) the University of Carabobo, Valencia, Venezuela; and (c) Cayetano Heredia University in Lima, Peru as case examples of locally governed biorepositories. Each of these entities had established human subject ethics approved biorepositories in response to Zika virus research needs with ability to share samples for advancing research and as reference materials. In the case of Colombia, they already had established the AEDES Network that included sample sharing within the network members. Their collective experiences are used to illustrate their biorepository decision-making governance and operational structures and how this infrastructure may set as examples of locally managed sample sharing mechanisms.

## Results

### Benefits survey

#### Characteristics of survey respondents

Forty-seven respondents completed the survey, and their replies were aggregated and analyzed. Respondents logged into the survey from Africa (n=15), North America (n=10) Europe (n=9), Latin America (n=9) and 5 were from the Asia-Pacific region (Fig 1). They represented a variety of organizations from research institutions (n=23), not-for-profit or government institutions (n=22), public health and clinical laboratories (n=9), biorepositories (n=5) and commercial diagnostics laboratories (n=3); a few respondents (n=4) did not identify their affiliation. Respondents were able to select more than one category, for example both research and government institution. The participants also represented a broad group of stakeholders with a variety of roles, including principal investigators, public health officials, institution leaders, and laboratory or biorepository managers.

**Figure 1:**
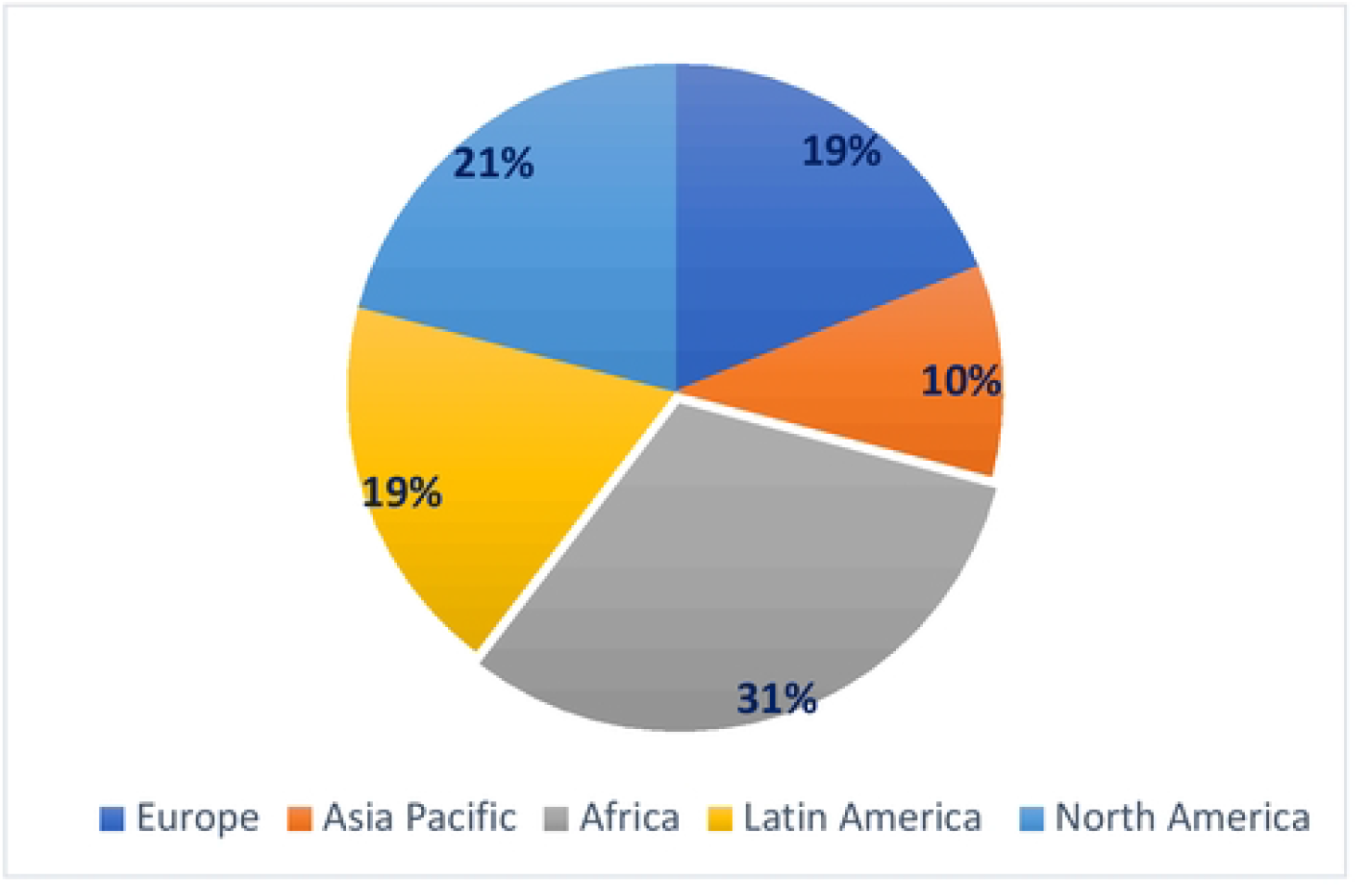
The geographic location of survey respondents.

#### Benefits of the VBS

Embedded in our 10-question survey (SI, Table 1) were 3 questions aimed specifically at understanding what respondents considered to be the key benefits for participating in a VBS. Survey respondents could choose all that applied to them from a list of potential benefits and rank their importance. These questions were: (a) which planned VBS benefits would be most important to you/your institution? (Q4), (b) which VBS support functions would be most valuable to your current biorepository or collection? (Q5) and (c) which additional resource would allow you to participate in the VBS (Q6).

The most important benefits for investigators or their institutions that ranked over 50% in favor of were: (a) the opportunity to network and collaborate with international partners (38/47, 81%), (b) capacity building for infectious disease specimen resources (35/47, 74%), (c) career growth and training opportunities (28/47, 60%), (d) greater visibility and utilization of specimens (26/47, 55%) and (e) additional funding opportunities 25/47, 53%) as shown in Table 1.

**Table 1.**
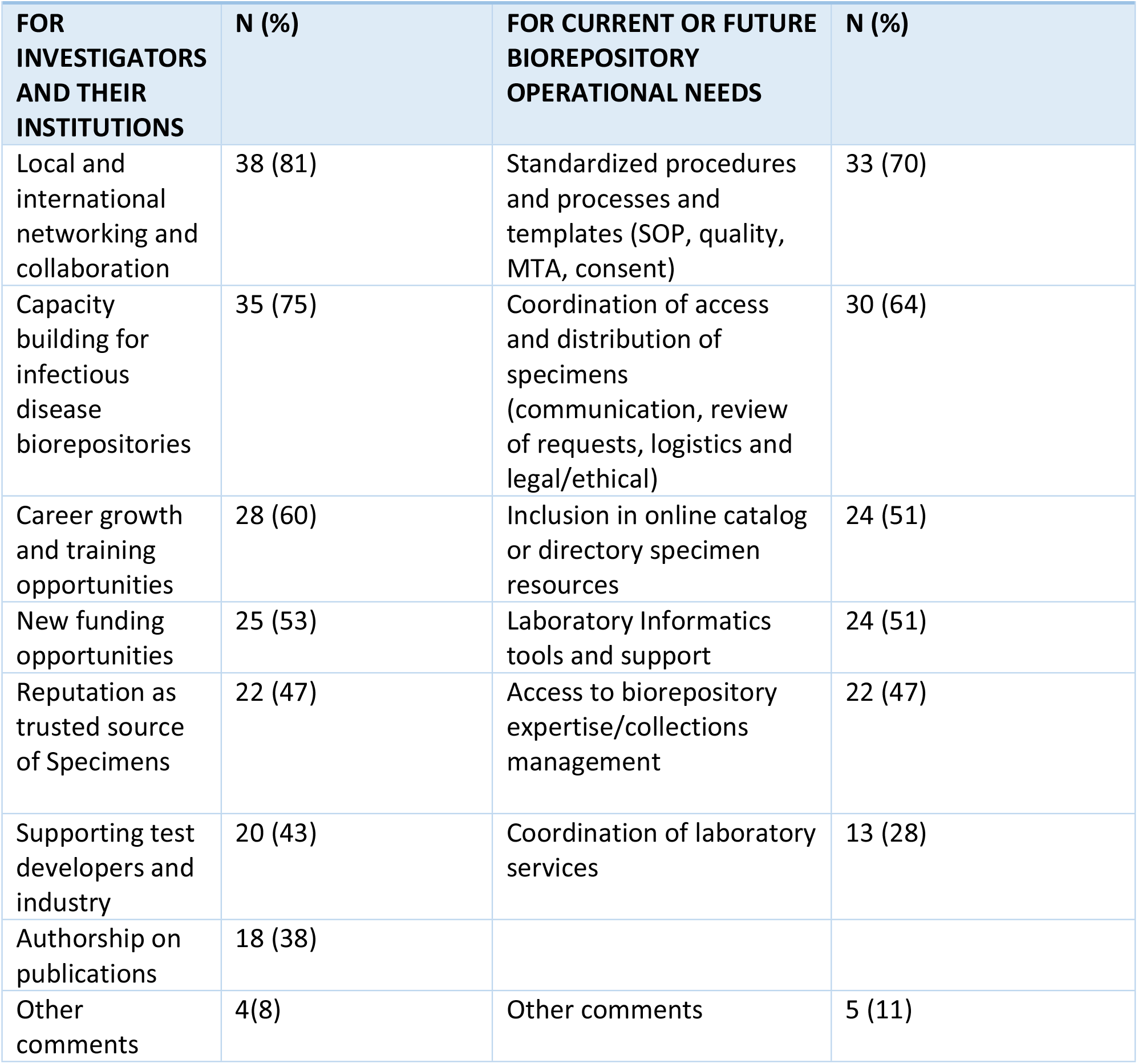
Virtual biorepository functions and services of greatest benefit listed by order of preference.

When survey respondents were asked to identify which features/functions of the VBS would be of highest benefit to their current or future local biorepository operations (Table 1), the most often selected functions were (a) the availability of standardized procedures and processes including Standard Operating Procedures (SOPs), quality standards, templates for Material Transfer Agreements (MTAs) and for consent procedures, (33/47, 70%); and (b) the coordination of logistics for accessing and distributing specimen, such as communications, review of requests and assistance with legal and ethical issues (30/47, 64%) with (c) inclusion in a catalog and directory and laboratory informatics tools and support being equally selected as valuable features by 24/47 (51%). The feature of a catalogue and directory was the top of list of valuable VBS function for diagnostics developers, especially smaller companies interviewed, as described in the next section.

In addition to the direct responses to the survey questions, an open comments section offered an opportunity to suggest additional desirable benefits provided by the VBS that were not included in the questions. The comments suggested that the VBS could: (a) serve as forum for sharing successful problem-solving approaches (e.g., legal issues), (b) support start-up of new biorepositories, (c) promote research and development capacity for preparedness in LMICs, and (d) ensure proper attribution for specimen providers. It was also recommended that the VBS should strive for a governance framework in which all partners are treated as equal members. In our questionnaire, we also asked what additional resources the investigators or institutions may need to participate in the VBS. Only 42/47 responded to this question and for the majority they needed all the resources while others were more explicit in needing expertise in biorepository/collections management, informatics support and tools, equipment and maintenance, staff and facility improvements. These results would suggest that the need for resources goes beyond expected financial needs and reflects specific needs for the operation of the VBS.

#### Barriers to sample sharing

A frequently invoked barrier for sharing samples across borders is a party’s compliance with the benefit sharing requirement of the Nagoya Protocol (NP). In Q7, respondents were asked whether their country had any policies in accordance with the NP and whether it would prevent them from contributing samples under the VBS. An interesting, but not surprising result was that more than half of the responses (53%, 25 out of 47) indicated lack of familiarity with their own country’s regulations regarding the NP and the respective impact on sample sharing. Respondents from countries with specific policies in accordance with the NP were asked to elaborate whether the rules would prevent sharing outside the country. Some (19%, n= 9) indicated that sharing was possible with approval from appropriate authoritative officials, such as their respective Ministries of Health. When international sharing was restricted, some believed they could still participate and use their local specimens to test in-country, if reagents or kits were provided.

#### Other findings

We asked respondents about the feasibility of contributing to the VBS a set of qualified specimens from their COVID-19 specimen collections to be used for creating evaluation panels and 16 /40 of the responses (40%) indicated that they could provide such specimens. We plan to follow up on this important question to explore whether the VBS could provide specimen panels as a service. Several additional items for follow up were indicated in comments (SI Table 1, Q10) including: (a) barriers to sharing such as shipping of specimens; recommendations for a successful partnership to be based on equity, respect and credit for contribution, (c) support for a VBS as a catalyst for research collaborations and, (d) the benefit of the VBS providing access to biorepository expertise.

Finally, 40/47 respondents replied via comments to Q8 (SI Table 1) which asked them if they would be inclined to participate in a VBS and why they would join: 29/40 indicated they would join and the reasons were about samples, research, networking and an opportunity for benefits and for research affirming the choices in Q4-Q6 (SI Table 1).

### Interview results with consumers and providers: identifying unmet needs and barriers to access

#### Characteristics of interviewees

We specifically invited representative organizations with different perspectives and needs for detailed interviews: (a) developers of in vitro diagnostics with differing market sizes and (b) organizations providing specimens and related services, reference sample providers and managers of networks of collections.

A total of 11 interviews were conducted, We were able to Interview 4 small companies and 2 large/midsize companies that marketed their products globally. For analysis, we made two groups: (a) 4 small companies and 2 large/midsize for-profit enterprises with global markets and, (b) 5 not-for-profit enterprises, with a company that provided external quality assessment samples and those that were academic but government supported or government organizations. Despite the small number of interviewees, we were able to capture a wide range of organizations that provided us key insights.

#### Gaps and barriers for access to specimens

Among those interviewed, we found notable differences in the priority of concerns among the organizations with some common gaps or barriers when we compared their replies, summarized in Table 2.

**Table 2.**
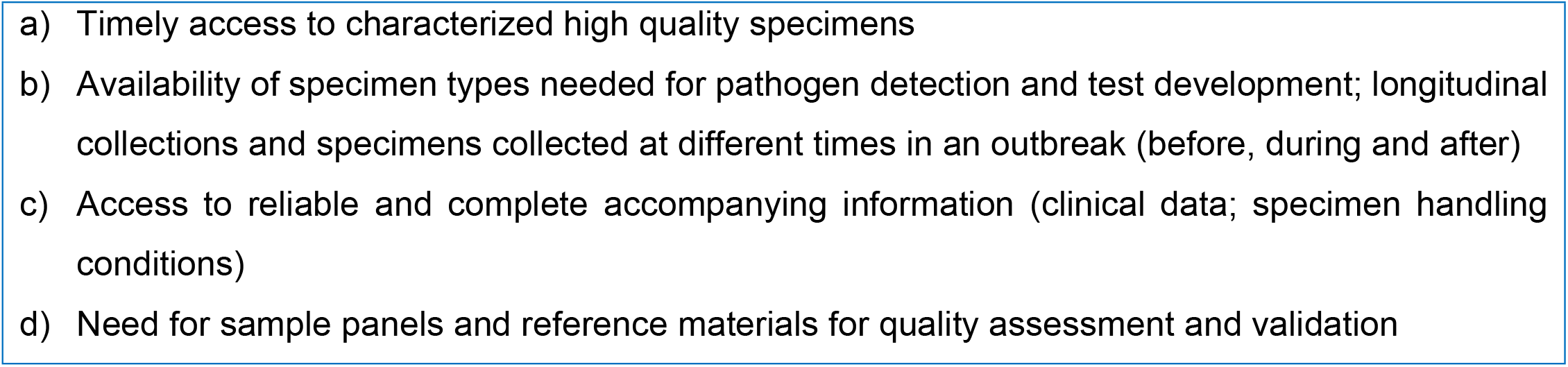
Common gaps and barriers of high concern for access to specimens.

A detailed summary of the interview results is included in the SI Table 2, and demonstrates the common concerns as well as differences in the types of barriers and hurdles of highest concern for different types of organization interviewed. Availability of some specimen types both in the context of COVID-19, but especially for the previous outbreaks (limited geographically, seasonally, recurrence, etc.) was a universally recognized concern by all interviewees.

Some differences in ranking of the barriers were also evident from the interviews (SI Table 2), depending on the type of organization represented. For commercial IVD developers, a comparison of larger vs small IVD developers indicated that for smaller companies identifying reliable trusted sourcing of specimens was a very high priority for accelerating time to development and approval of new diagnostics. Interviews indicated that such companies need to identify and negotiate with new sources of specimens for each new pathogen or obtain specimens from commercial sources, while larger companies are more likely to have long-standing collaborations with existing legal agreements. Related to sourcing was also access to reliable and sufficient data to meet regulatory requirements, as such data often is inadequate for commercially acquired specimens and of convenient use of left-over specimens whose control and oversight are not strictly managed. Data access restrictions, especially in European counties, due to privacy rules may also impact access for all specimen users. Cost of specimens was also cited as a significant financial burden. These issues affected all types of organizations but appeared to disproportionately impact small companies and was more of a concern early in outbreaks resulting in considerable delays and lost opportunity for product development.

Organizations that work in the international sphere, primarily the not-for-profit organizations that broker specimens for research and commercial and other entities, identified the Nagoya Protocol and related policies as the greatest barrier to access. They also expressed concern about the complexity and disparity in regulations between countries regarding export and import of biospecimens and biosafety/biosecurity, that greatly impact efficient timely sharing of specimens. Lack of trust was identified as important barrier for accessing specimens from LMICs, including ethical concerns about appropriate use of specimens and a need for greater awareness and investment in capacity building, training and emphasis on benefits for partners from LMICs. This group also provided specific recommendations regarding features of the VB that in their experience would meet the needs of various stakeholders (Table 3), specifically these recommendations aimed to reduce the complexities around sample access and specimen sharing under an operational structure to maintain quality and confidence in the materials to be accessed.

**Table 3.**
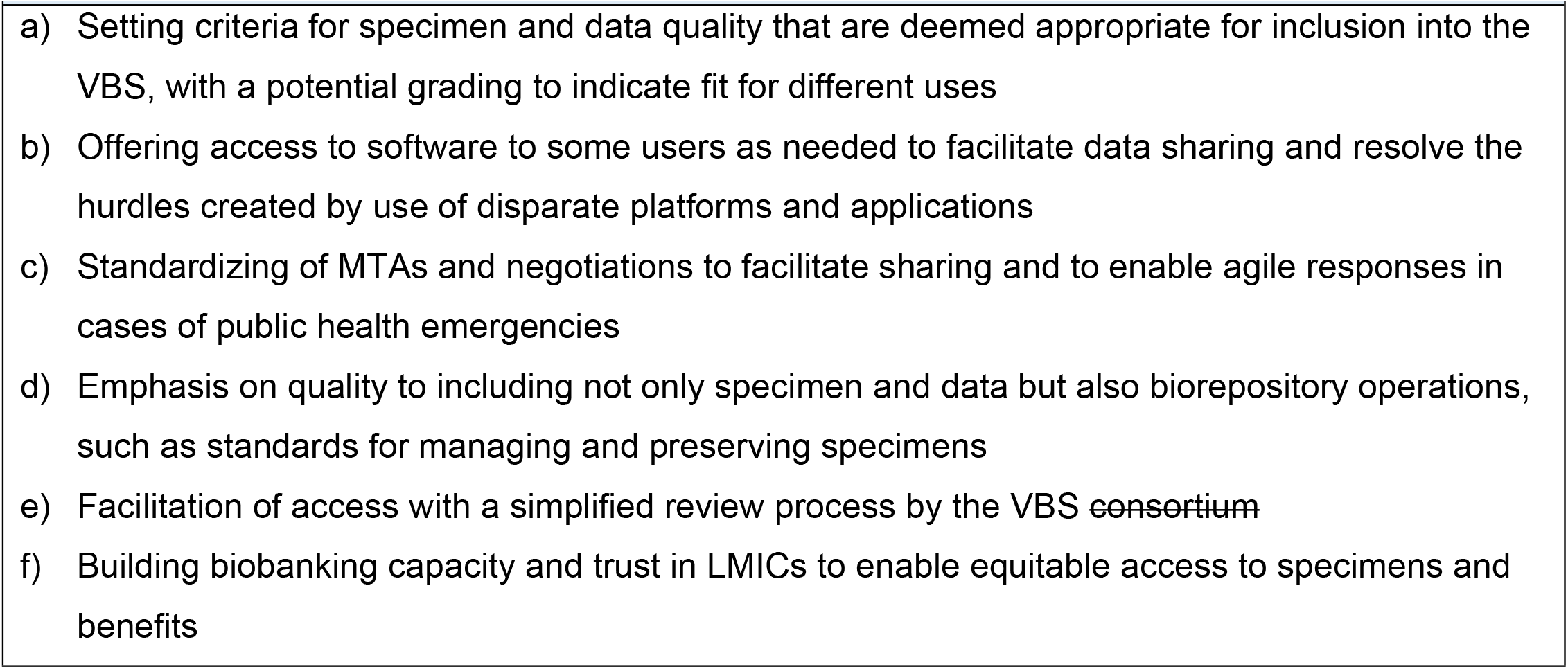
Recommendations regarding features of a VBS.

### Local governance models to inform the VBS framework

Here we describe locally-managed sample sharing model to inform how VBS could be structured. During the Zika epidemic in South America (2016), ZIKAlliance partner institutions developed a common governance structure for local sample sharing to facilitate and to provide access to samples for research studies (Figure 2). The oversight research plan and overarching ethics are determined in the ZIKAlliance framework and at each site, with the decision and access to specimens residing with the local principal investigator (PI) and the local Selection Panel (SP). Samples for sharing are listed in the public-facing catalogue with requests made through the local PI or leadership team and then reviewed by the SP for its ethical intended use and research value. When a request is approved, the PI the forwards it for to the next level to the regulator, which in Venezuela and Colombia is the Ministry of Health and Social Protection and in Peru the Ministry of Health (SI Figures 1 A-C). In a specific case of sample transfer from Colombia to France within ZIKAlliance, a submitted request obtained the requisite local IRB approval with the appropriate scientific justification for the transfer of the samples - namely in this case that the inclusion of a new molecular test to be conducted in a central laboratory in alignment with the primary objectives of the ZIKAlliance study. So following local approval, the sample transfer was then submitted for permission to share with the Ministry of Health and Social Protection of Colombia (https://www.minsalud.gov.co/Paginas/default.aspx) based on the Resolution 8430 of 1993 and Article 5 of the Resolution 3823 of 1997. The approval process, from time of request to the PI can conclude within 1-2 months with official notification from the Ministry of Health to the PI.

**Figure 2.**
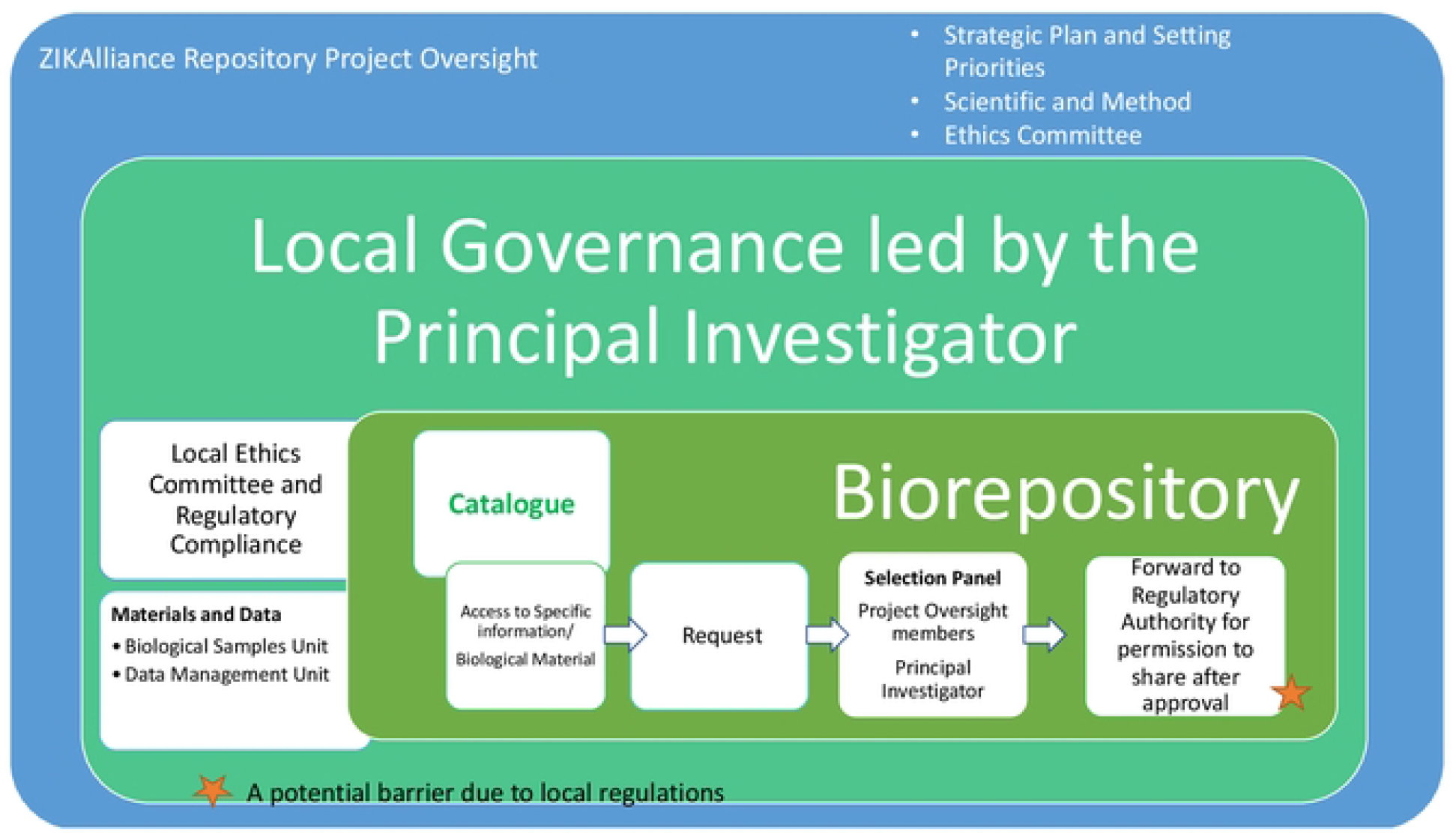
Governance structure of the ZIKAlliance local biorepositories.

Another example linked to this effort is the AEDES network in Colombia, where a Scientific Management Committee is responsible for coordinating the interaction and monitoring the progress of the technical, scientific and institutional strengthening components of the network in accordance with its Scientific Advisory Board and their Institutional Review Board or Ethics Committee.

In most of these projects, a publicly shared catalogue of the biological specimens is envisaged as the starting point (Figure 2) and the processes developed in the governance structure, allowed for a careful, thoughtful process at the hand of the local PI for sharing samples. This effort also then allows for recruitment and replenishment of samples. A future challenge beyond ZIKAlliance will be the availability of funds and scientific resolve to maintain this system.

## Discussion

### Benefits survey results and its implications

Overall, our results show that networking opportunities and capacity building for infectious disease biorepositories were considered of high importance, as confirmed also in the open comments section of the survey. The responses suggested areas for further follow up that we intend to pursue through a Delphi-prioritization exercise. In order to operationalize the VBS, we will like greater clarity on what could constitute capacity building, the implications of the Nagoya Protocol on specimen sharing and how different countries have adopted benefit sharing recommendations [19]

The key features of the VBS would be its role as ‘trusted broker’ or “navigator” for guaranteeing the source of high-quality specimens for future pandemic threat agents, which includes the availability of granular information on the clinical phenotype linked to the sample and the potential of sequential samples over the course of the illness. The broker status also includes enhanced accessibility due to successful navigation of legal and logistics hurdles for which good examples already exist.

Another area of discussion is the use of the VBS as a source of specimens for reference materials and panels. For example, one of the models considered for the VBS, is to request specimen providers to identify and set aside a few large volume well-characterized high-quality specimens that can be accessed through the VBS, for use for in evaluation of reference panels needed for calibration of diagnostics; this would require setting aside only a portion of their respective collections rather than their entire biorepository or specimen collection for a pathogen of interest. For this to happen, the local sites are responsible to ensure their local IRB approvals and informed consent processes clearly address the use of the set-aside samples. About 40% of respondents indicated that they would consider setting aside a small set of qualified specimens from their COVID-19 specimen collections for sharing within the VBS. Most respondents indicated the need for follow up with more information.

### Interviews with specimen consumers: identifying unmet needs and barriers to access

In the interviews, many of the participants from various types of organizations voiced the hope that the features of the VBS would provide solutions to barriers identified, differing primarily in ranking of priorities based on the types of organizations and their mission, such as small compared to large established diagnostics developers. The key features of this VBS identified by all were: (a) serving as a trusted, reliable resource for identifying and accessing characterized specimens and associated data and, (b) availability of characterized specimens of sufficient volume and quality fit for evaluation panels. Characterization of the samples in a standardized process will have benefits partners especially in LMICs where there is less opportunity to conduct the characterization themselves, thus providing them a resource that could allow them to develop diagnostics locally for their specific market. An example of potential benefit that the respondents agreed is a cross collaboration to create clinical evaluation panels to validate and utilize for license submission for diagnostic test kits, especially where public health and test developers can provide access to clinically confirmed case samples that would be difficult to assemble by a single entity, such as the comprehensive panel of sera from patients with various states of Lyme disease and healthy persons .for Lyme disease serological diagnostic testing [23].

### Comparison of local governance models

The local governance models were driven by the idea of local management of specimens. In this case, the consortium representatives review all applications, but final decision making about sharing is still retained by the institution who has the local control of the specimens.

By promoting local biorepositories in LMICs, we offer technology transfer opportunities and capacity building even if dedicated funding cannot be guaranteed. We need to put emphasis on fit-for-purpose technologies appropriate for LMIC settings (lyophilized and dried stocks for example instead of low temp freezing). There needs to be high transparency standards because of the sensitivity of partners in LMICs about misuse of specimens. The focus on local biorepositories echoes the need expressed in the interviews for help with the operational challenges, including harmonized platforms and logistics.

This can be implemented by formal agreements between partners in the system, with the selection of partners based on willingness to adhere to the governance criteria (specimens, data, biorepository operations, MTA, sharing etc.). Such services will need constant monitoring, adapting to stakeholders and specifically user’s needs.

### Limitations of this study

The survey was delayed from conception in 2019 to completion in 2021, interrupted by the on-going COVID-19 pandemic. We made attempts to enroll all who had expressed interest through our workshops to participate in our survey and interview on the concept of a VBS. All the respondents expressed interest in solving existing gaps for specimens’ access, but not everyone had a complete or in-depth understanding of all the issues around sharing or the requirements of maintaining a non-centralized, federated biorepository. An outcome of this first survey is to understand the type of engagement we might expect in a proposed VBS operation and we are actively pursuing steps to operationalize the proposed VBS.

## CONCLUSIONS

Biorepositories, as most in biological medical sciences professionals understand it, are narrowly focused collected for research on specific conditions (cancer, precision medicine, genetic and metabolic diseases, as examples) and more recently, for digital data. They typically require major investments in infrastructure and consistent long-term support, such requirements have then relegated biorepositories to be primarily established in higher income countries rather than countries with fewer resources where new pathogens may emerge. A global unmet need has been access to quality specimens that represent the diversity of where a disease “X” may yet emerge from. In the absence of specimens from low resource settings, laboratory tests and vaccines may be developed without the opportunity to fully assess their performance in all populations.

The vast spread of the COVID-19 pandemic and its duration has provided an abundance of specimens--more than 100 sites identified by global search have identified themselves as COVID biobanks [1,14,16,20,24] but most have uncertain remit to what comes next--thus this dilemma offers an excellent opportunity to address challenges for storage, access, sharing and sustainability.

In our study, we included the perspective of the ‘clients’ who are interested in high quality specimens related to infections with outbreak potential. This allowed us from the specimen collection perspective to examine the needs and identify the barriers in sourcing specimens along with understanding the types of equitable benefits that participants themselves positively identify as acceptable. We examined what might constitute a fair exchange between the investigators collecting well-characterized specimens in LMIC and their clients; a system in which specimens of high value can be collected and replenished while benefits of recognition and research participation activities can be fostered.

In recent years, recognizing the lack of access and knowledge of types of specimens, multiple efforts have been set up to provide materials and have functioned successfully (EVA-G, ZIKAlliance, PATH, FIND) but they do not cover the entire spectrum of needs as evident by multiple calls for better collections and sharing efforts [1,5,17], we think this is where a VBS model could align and contribute. We especially recognize that there is a need for well-annotated collection of serum/plasma from diverse geographical and disease exposure experiences [14]. A focus on this type of specimens would allow us to build the required infrastructure and activities of the VBS as proof of concept.

We also recognize some national laws and recommendations preclude exporting specimens but partners in those situations are open to collaborative research; we think then, depending on the research or project, diagnostic kits and protocols can be shared and shipped along with reference calibration materials to the site for evaluation thus allowing for active research collaboration.

Biorepositories are of many types and function [7,25-27]. Here we explicitly focus on their contribution to global preparedness by enabling and accelerating research and development of interventions through rapid sharing of well annotated specimens to inform outbreak control strategies. Specimen collection and storage would need to be distributed over a diverse range of geography, ecozones, and demographics. Our idea of creating a distributed, grassroots biorepository is in alignment with concurrent efforts of the DxConnect and modeled after the successful virus exchange by EVA-G [2]. Our effort through TGHN and partnership with ZIKAlliance, ReCoDID and CREID allows us to fill in gaps in collections from geographically diverse ecozones and human exposomes. A virtual biorepository that functions as a trusted broker under a federated system on a demand-and-supply model could allow many to participate without high costs. Such a system would also allow local generators of biospecimens to receive benefits for setting aside potentially high value specimens. Sets of well-characterized and richly annotated samples can be queried in a database under a common governance framework would allow contributors and users alike to access materials respectfully as equal partners. Recent post-public health emergency epidemic reviews on lessons-learned for the future, all stated that preparedness must include access to data and specimens [2,6,28-31], yet support for biorepository functions remain as an unfunded mandate for many so the recommended actions will need champions to promote, socialize the benefits of specimen access and encourage investment in the sharing systems.

In this paper, we present arguments for building a trusted sample sharing system based on a distributed stewardship at the local level that manage the access to quality, well-characterized specimens for outbreak-prone infectious diseases as a global public good. Accessing specimens, even within established networks and projects, remain highly fragmented with procedures that often are not transparent and hampered by barriers leading to long lag time and can be very costly. We obtained knowledge, insights and advice through our workshops, questionnaires, interviews and case examples from public health experts, infectious disease researchers, biorepository managers, clinicians, policy makers, regulators, and especially diagnostic industry representatives to ascertain what might be appropriate entry points to better organize fair and equitable sample sharing experience that allows more engagement with source providers and stimulate research. The following challenges remain - (a) generating quality specimens, (b) overcoming barriers to accessing specimens (regulatory, cost, sharing mechanisms), (c) generate a supportive exchange infrastructure, define benefit packages, and finally, (e) maintain trust between parties; all add up to requiring the seeker of specimens to have persistence, commitment of precious time and financial support as the outbreak of interest slips away and then only to repeat this cycle again for the next outbreak. We propose a coordinated approach with logistics support and assistance with procedures to facilitate exchange between sample providers and user. This proposed system may work well and complement existing biorepositories efforts to provide an alternate way to shorten the time to obtain results for decision making for preparedness and response to future public health emergencies.

We want to propose a solution that maintains respect for contributors and users of biological materials. The concepts of equitable benefits sharing and capacity building are essential if our effort is to be successful and long lasting. And finally, a coalition of the willing will be needed to ensure funding, infrastructure and services to realize our vision. We look forward to formalizing the framework and operationalize the VBS globally.

## Data Availability

all data will be available

## Acknowledgements

The work was partially funded by the European Union Horizon 2020 Research and Innovation Programme under Grant Agreement No. 825746, and is supported by the Canadian Institutes of Health Research, Institute of Genetics (CIHR-IG) under Grant Agreement No. 01886-000. We are indebted to FIND (Stefano Ongarello and Dominique Allen) for co-sponsoring our first workshop and for joint discussions with PATH (Roger Peck and Helen Storey) on the concept of the VBS. We are grateful to Zainab Al-Rawni, Adam Dale and Trudie Lang of TGHN for hosting our VBS website and access to the workshops.

## Author Contributions

Conceptualization: Judith Giri, Thomas Jaenisch, May Chu,

Survey development and analysis: Judith Giri

Visualization: Judith Giri, Laura Pezzi, Rosa Margarita Gèlvez Ramirez, Rodrigo Cachay

Writing-original draft: Judith Giri, Thomas Jaenisch, May Chu

Writing- review and editing: All authors

## SUPPORTIVE INFORMATION (SI)

**Figures and Tables**

### Figures

SI, Figure 1. Logistical diagrams on the governance of locally managed biorepositories

**Figure 1A:**
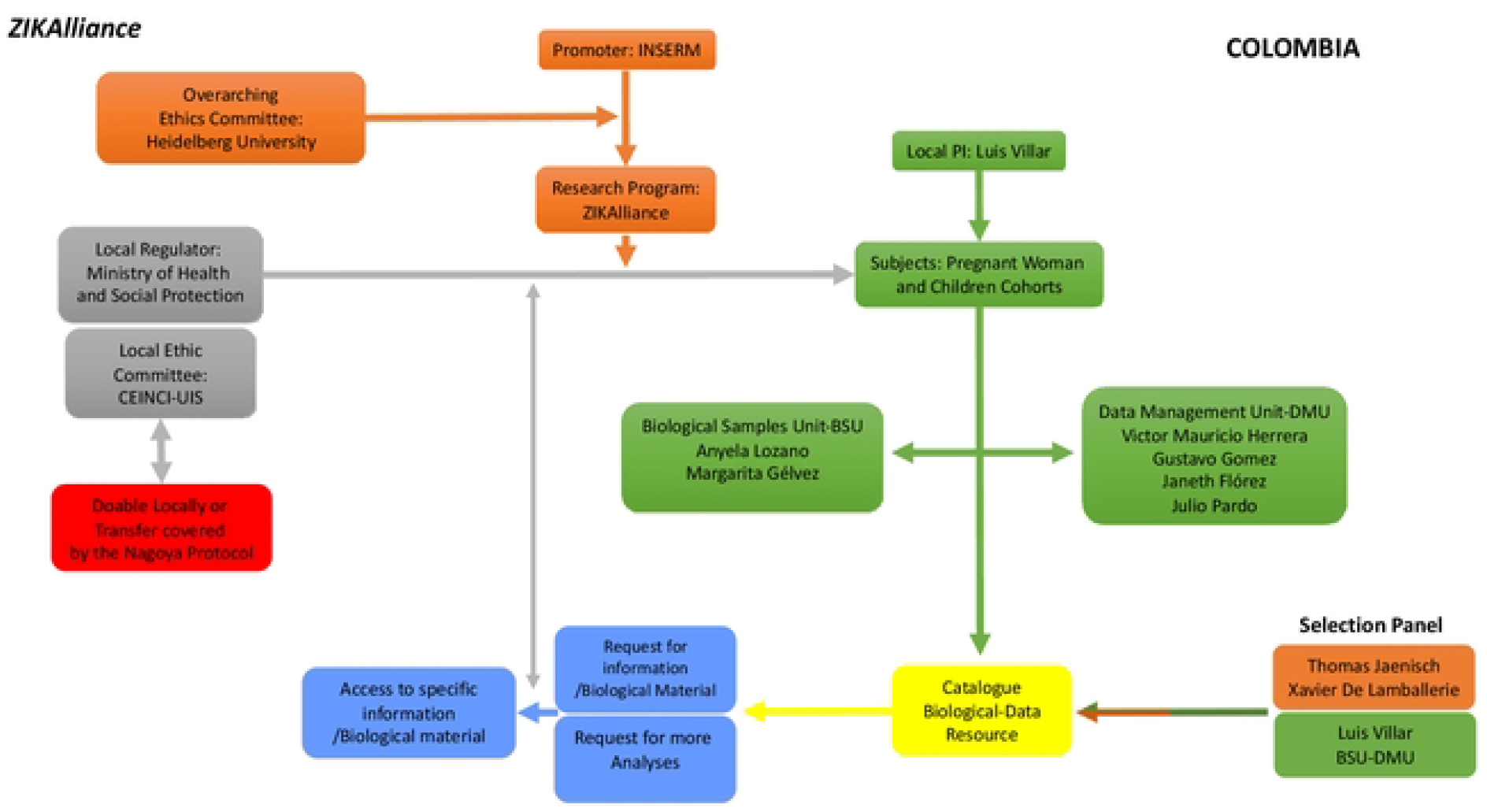
The Industrial University of Santander, Bucamaranga, Colombia and the Fundaciôn INFOVIDA and Centro de Atención y Diagnóstico de Enfermedades Infecciosas-C

**Figure 1B:**
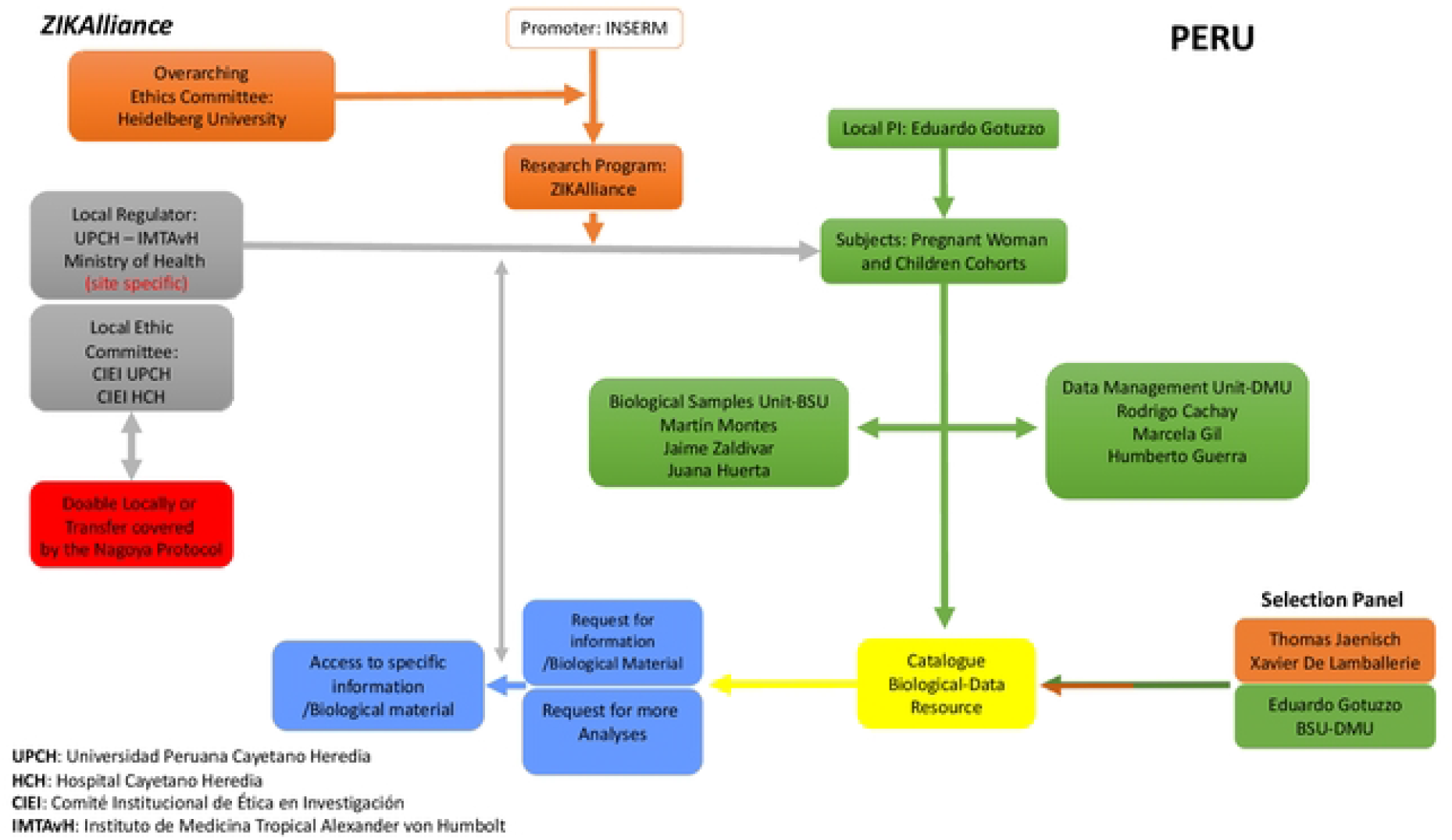
Cayetano Heredia University, Lima, Peru

**Figure 1C:**
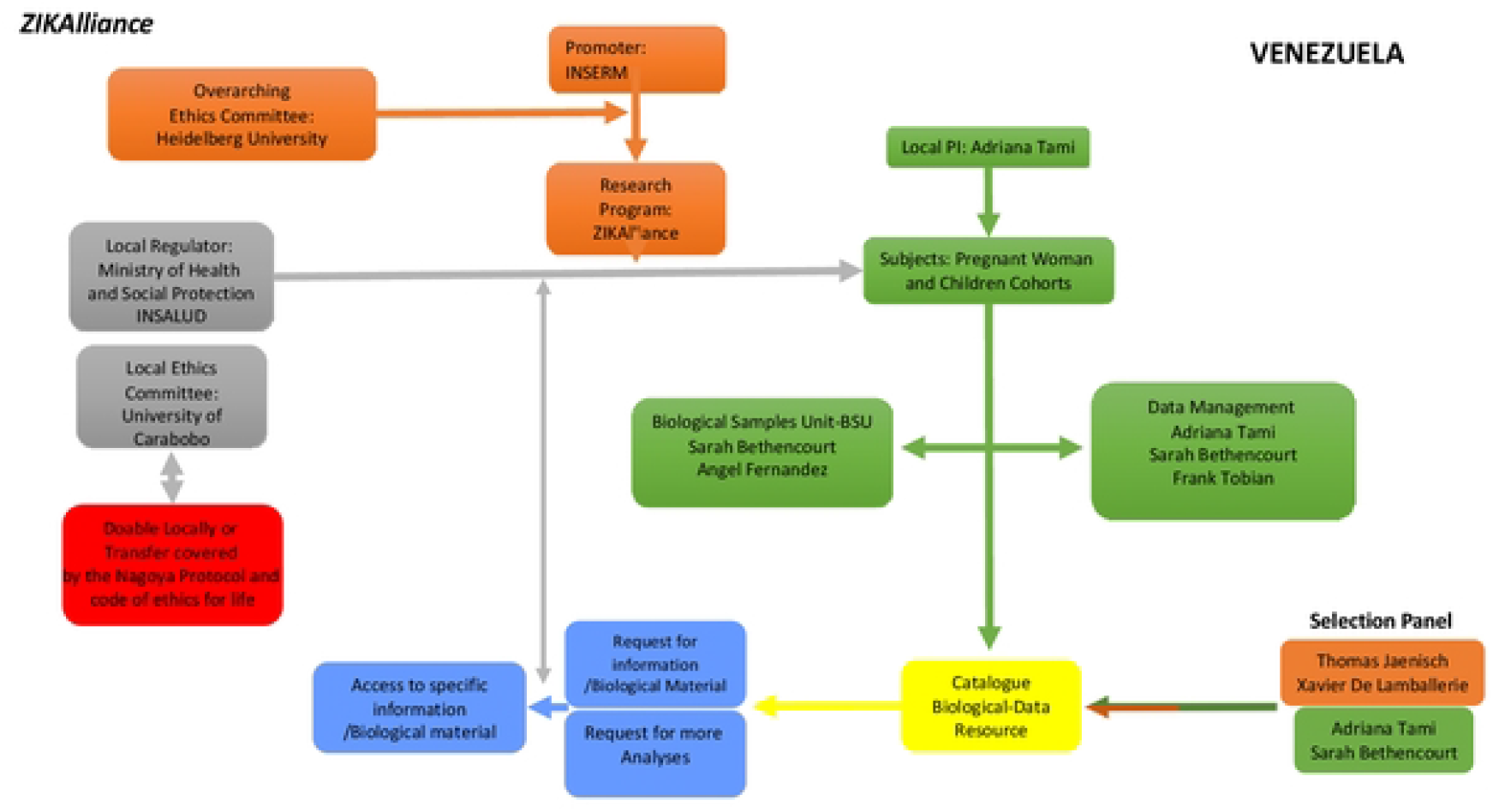
University of Carabobo, Valencia, Venezuela

### TABLES

**SI, Table 1:** Virtual Biorepository (VBR) Benefits Questionnaire.

| Table 1: Virtual Biorepository (VBR) Benefits Questionnaire |  |
| --- | --- |
| CATEGORY | QUESTIONS |
| Profile of respondents | 1. What is the nature of your institution |
|  | 2. What is your role |
|  | 3. What is your geographic location |
| Benefits of participation: support functions and resources | 4. Which planned VBR benefits would be most important to you/your institution? |
|  | 5. Which VBR support functions would be most valuable to your current biorepository or collection? |
|  | 6. Which additional resources would allow you to participate in the VBR? |
| Benefit sharing and the Nagoya protocol | 7. Does your country have a specific policy in accord with the NP? |
|  | - If yes, would it prevent you from sharing specimens outside the country? |
|  | - If yes, would you be willing to test in country? |
| Interest in participation | 8. Would you be interested in participating in a VBR |
| Contributing to reference material and panels | 9. Would you be willing to set aside limited numbers of samples for use as reference materials and as part of evaluation panels? |
| Other benefits? | 10. Comments |

SI, Table 2. Interviews to assess needs and barriers to accessing specimens

**SI, Table 2.** Interviews to assess needs and barriers to accessing specimens.

|  |
| --- |
| **1. What are your greatest hurdles for access to specimens?**<br>*Examples of issues to consider:* identifying sources; reliability of sources; cost; quality of specimens; completeness and reliability of accompanying data; legal hurdles: Material Transfer Agreements (MTA) and negotiations; other |
| **2. What are your current strategies for access?** |
| **3. What type of specimens have been difficult to source for COVID/19 and other infectious diseases?** |
| **4. What are some of the challenges for access to specimens in LMICs?** |
| **5. Which features of a Virtual Biorepository would be most useful?**<br>*VB features to consider:* coordinated, one stop access to specimens; an online catalog or directory of sources; more affordable specimen sources; trusted source of qualified specimens; facilitated access including MTA negotiations and logistics; other |
| **6. Would you be willing to contribute to creation of a manufacturer's panel <sup>(1)</sup>** |
| **7. Do you have any additional comments and recommendations?** |

SI, Table 3

**SI, Table 3.**
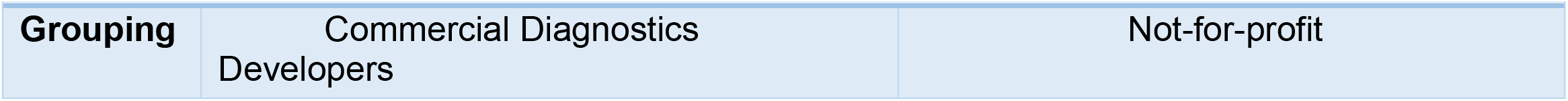
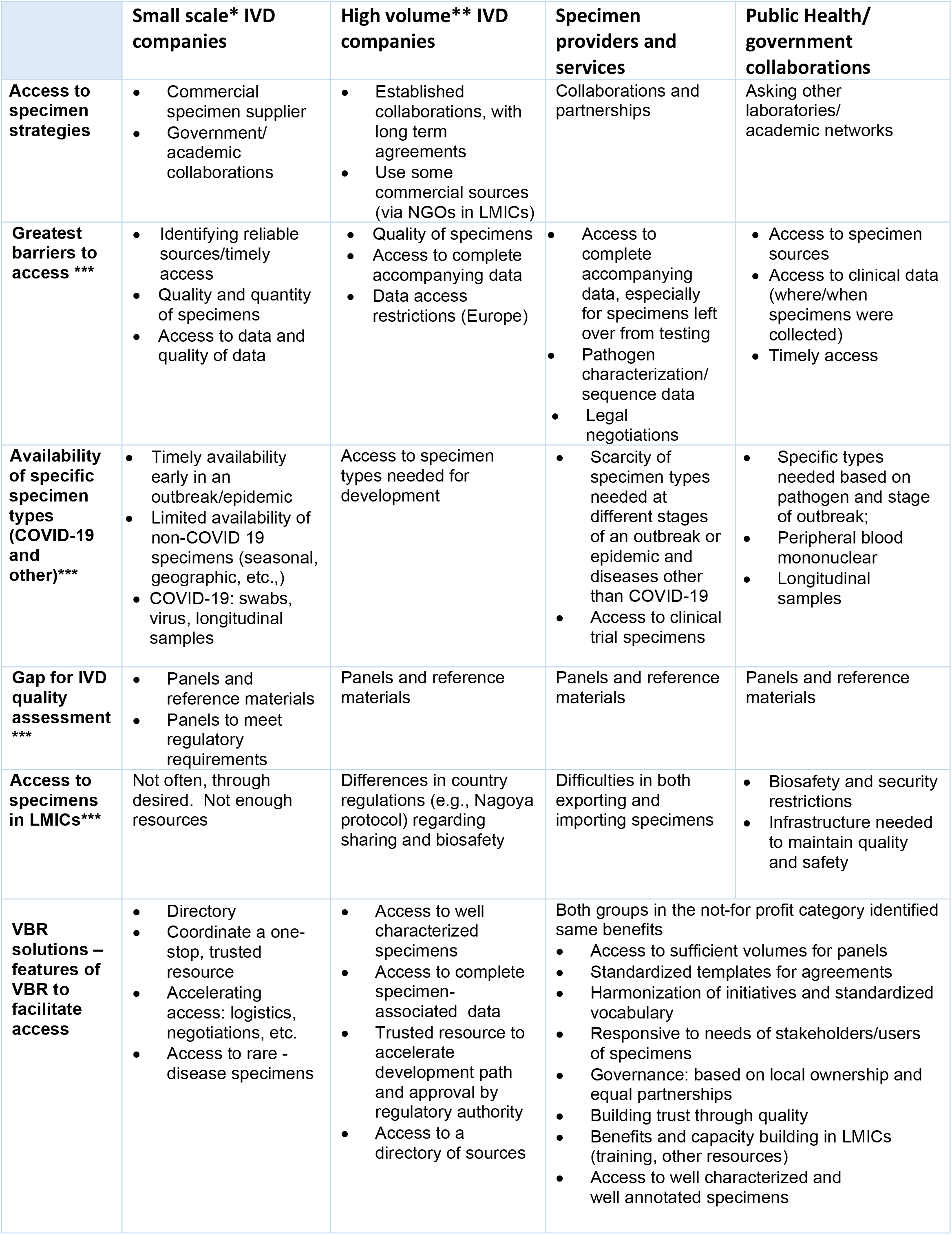
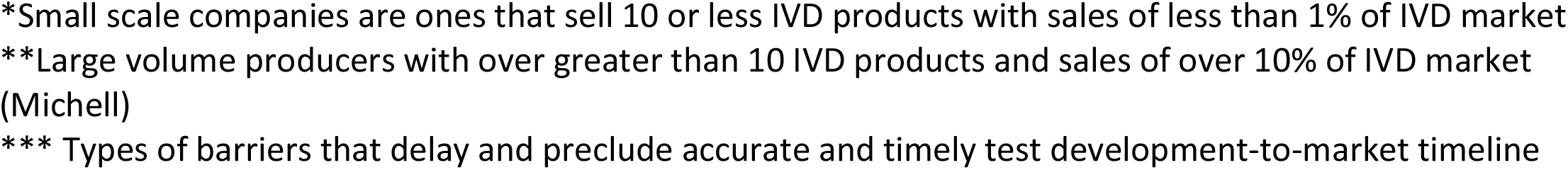
Interview summary of gaps and barriers to access.

